# Exploring the Perspectives of Clients and Clinicians Regarding Digitally Delivered Psychotherapies Utilized for Trauma-Affected Populations

**DOI:** 10.1101/2024.04.09.24305560

**Authors:** Sidney Yap, Rashell Wozniak, Katherine Bright, Matthew RG Brown, Lisa Burback, Jake Hayward, Olga Winkler, Kristopher Wells, Chelsea Jones, Phillip R. Sevigny, Megan McElheran, Keith Zukiwski, Andrew J Greenshaw, Suzette Brémault-Phillips

**Affiliations:** Department of Psychiatry, Faculty of Medicine and Dentistry, University of Alberta, Edmonton, AB, Canada; School of Clinical Child Psychology, Faculty of Education, University of Alberta, Edmonton, AB, Canada; Heroes in Mind, Advocacy, and Research Consortium, Faculty of Rehabilitation Medicine, University of Alberta, Edmonton, AB, Canada; Department of Computing Science, Faculty of Science, University of Alberta, Edmonton, AB, Canada; Department of Emergency Medicine, Faculty of Medicine and Dentistry, University of Alberta, Edmonton, AB, Canada; Department of Child and Youth Care, Faculty of Health and Community Studies, MacEwan University, Edmonton, Alberta, Canada; Department of Occupational Therapy, Faculty of Rehabilitation Medicine, University of Alberta, Edmonton, AB, Canada; Faculty of Education, University of Alberta, Edmonton, AB, Canada; Wayfound Mental Health Group, Calgary, AB, Canada; Haikei Health, Edmonton, AB, Canada

**Keywords:** Web Based Intervention, Psychotherapy, Access to Therapies, Trauma, Trauma-focused Psychotherapy, Health Services, Military, Veteran, Public Safety Personnel, Implementation Science

## Abstract

During the COVID-19 pandemic, many clinical sites shifted towards digital delivery of mental health services. However, there is still much to learn regarding using digitally delivered psychotherapies in trauma-affected populations, including military members, Veterans, and public safety personnel. This study examined perceptions of psychotherapies utilized for trauma-maffected populations, as reported by Canadian military members, Veterans, and public safety personnel who completed such interventions and mental health clinicians who provided them. Specifically, we explored the imposed shift to digital health use, what changed with this rapid shift, what needs, problems, and solutions arose, and important future considerations associated with delivering trauma-focused and adjunct treatments digitally.

Quantitative survey data were collected from 11 Canadian patients (military members, Veterans, and public safety personnel with post-traumatic stress injury) and 12 Canadian mental health clinicians. Survey questions were adapted from the Alberta Quality Matrix for Health (AQMH) and Unified Theory of Acceptance and Use of Technology (UTAUT) model. As a follow-up, participants were invited to participate in either a semi-structured qualitative interview or focus group to further explore their perspectives on digitally delivered trauma-focused and adjunct therapies. Four clients and 19 clinician participants participated in an interview or focus group.

In survey and interview/focus group results, patient and clinician participants reported that digitally delivered trauma and adjunct therapies offered similar treatment effectiveness as in-person delivery while also improving treatment access. Participants indicated unique advantages of digital delivery, including the increased accessibility of treatment, cost effectiveness, and more efficient use of resources. However, some participants struggled with using digital platforms and felt less comfortable working in a digital environment. Further research with a larger, more diverse population is required to corroborate our results and identify other avenues in which psychotherapies utilized for trauma-affected populations can be engaged with and improved upon.

**Author Summary:** Many mental health service sites were faced with rapid and unexpected shifts towards digital delivery of mental health services to comply with mandated physical distancing restrictions put in place during the COVID-19 pandemic. There is still much to learn regarding using digitally delivered psychotherapies in trauma-affected populations, including military members, Veterans, and public safety personnel. This study examined perceptions of Canadian military members, Veterans, and public safety personnel who completed, and mental health clinicians who provided, psychotherapies utilized for trauma-affected populations. This exploration aims to increase our understanding of the strengths and limitations of this mode of delivery. Patient and clinician participants reported that psychotherapies for trauma-affected populations offered similar treatment effectiveness as in-person delivery, while also improving treatment access. Participants indicated unique advantages of digital delivery, including increased accessibility of treatment, cost effectiveness, and more efficient use of resources. Some participants reported struggling with the use of, and felt less comfortable working on, digital platforms. Further research with larger, more diverse populations is required to confirm our results and identify other avenues for using, and improving on, psychotherapies for trauma-affected populations.

## 1. Introduction

The COVID-19 pandemic substantially impacted mental health and approaches to service delivery. In Canadian poll results, over 40% of individuals reported experiencing mental distress early in 2021 when COVID-19 related precautions were commonplace (Statistics Canada, 2021). The pandemic further stressed groups at high risk for potentially psychologically traumatic event exposures (PPTEs, e.g., exposure to actual or threatened death, serious injury, or sexual violence) (APA, 2022; Heber et al., 2023). Such groups include public safety personnel (PSP) and frontline healthcare workers, as well as currently serving military members and Veterans (Carleton et al., 2018; Heber et al., 2023). PPTEs can have physical impacts (e.g., digestive distress, cardiovascular disease), result in behavioral challenges (e.g., maladaptive responses like burnout, alcoholism, suicidality), and induce post-traumatic stress injuries (PTSI, e.g., post-traumatic stress disorder [PTSD], major depressive disorder) (APA, 2022).

Even prior to the pandemic, trauma-affected populations like PSP, Veterans and military members experienced a higher risk of mental health difficulties. A pre-COVID-19 survey found that 44.5% of Canadian PSP self-reported a mental disorder diagnosis (Carleton et al., 2018). Further, 23.2% of this Canadian PSP population yielded a positive screen for PTSD (Carleton et al., 2018). Public Safety Personnel also exhibit elevated psychopathology despite prior treatment, indicating clinical complexity and the potential need for transdiagnostic, specialized, or ongoing mental health support (McCall et al., 2021). Canadian Armed Forces (CAF) Regular Force Veterans exhibited similar prevalence for PTSD (16%) as PSP. Additionally, this population reported mood disorder prevalence of 21% and anxiety disorder prevalence of 15% (Van Til et al, 2016).

These unique mental health challenges may have been compounded by the COVID-19 pandemic (Clemente-Suarez et al., 2021). According to the Public Health Agency of Canada, those who met criteria for PTSD during the pandemic were more likely to report being impacted by the pandemic (and mandatory public health restrictions and protocols) in terms of physical health problems, difficulties in meeting financial obligations/essential needs, and facing challenges in personal relationships with household members. These individuals were also more likely to report symptoms of anxiety and depression within the past two weeks, greater lifetime suicidal thoughts, and increased use of alcohol and cannabis (Public Health Agency of Canada, 2021).

Transformation in service delivery was prompted by the escalation of mental health concerns due to COVID-19 and the need to adhere to physical distancing mandates and government restrictions. While utilizing digital modalities to deliver psychotherapeutic interventions was not a novel innovation when the COVID-19 pandemic began (see Gros et al., 2018 for a review), the pandemic required an unprecedented rapid transition from in-person to digital methods (e.g., teletherapy, telemedicine, eHealth, mobile health) (Torous & Keshavan, 2020). Considering mental health consequences of the COVID-19 pandemic (Statistics Canada, 2021; WHO, 2022), digital mental health interventions (DMHI) offered a cost-effective alternative to in-person mental health treatment while complying with public health requirements for physical distancing during the pandemic (Philippe et al, 2022). Such interventions allowed for access to timely and secure trauma therapies that are critical to supporting the transdiagnostic needs of PSP, military members, and Veterans (Smith-MacDonald, 2020). Accordingly, the COVID-19 pandemic greatly accelerated uptake of DMHI (Bautista & Schueller, 2022).

Research indicates that digitally delivered Prolonged Exposure (PE), Eye Movement Desensitization and Reprocessing (EMDR) and Trauma-Focused Cognitive Behavioral Therapy (TF-CBT) may be effective at reducing PTSD symptomatology (Jones et al., 2020; Perri et al., 2021). DMHI generally provides clients with increased convenience, comfort, and access to treatment (APA, 2015; Titov et al., 2019). Increased access to care can lead to substantial cost and time savings for clients (Philippe et al., 2022). DMHI may also aid in decreasing stigma related to accessing mental health care (Lattie et al., 2022).

Despite promising findings supporting the use of DMHI for general mental health care, evidence supporting the wide scale use of DMHI for clients experiencing PTSI is comparatively scarce. Few randomized controlled trials (RCTs) have directly compared the effectiveness of digitally delivered and in-person trauma interventions (Fleuty & Almond, 2020; Jones et al., 2020). Some Veteran clients reported that DMHI were negatively impacted due to Internet connection issues and because therapy delivery felt distant and impersonal in the digital environment (Fleuty & Almond, 2020). Those with greater symptom severity may also be less likely to complete treatment programs delivered digitally, as reported in a recent study evaluating the use of Internet-Delivered Cognitive Behavioral Therapy (ICBT) in PSP (Beahm et al., 2021). There is a clear need to explore mitigation of potential risks and disadvantages of using digital trauma interventions for those experiencing PTSI, including appropriate assessment of client compatibility, obtaining consent in a secure manner, ensuring safety in the context of suicidal ideation or behavior, adapting therapy protocol to the digital environment, and ensuring ongoing client privacy (Jones et al., 2020).

Clinician experiences and comfort with delivering DMHI can also be an important contributor to treatment success. Most clinicians supporting those with mental health challenges did not have prior experience providing digital services, trauma-specific or otherwise, prior to the COVID-19 pandemic (Békés et al., 2021; Aafjes-van Doorn et al., 2022). Clinicians have reported many concerns with providing DMHI, including a lack of emotional connection with patients, increased distraction (for both client and clinician), concerns with privacy and confidentiality, and clinician work-life balance (Békés et al., 2021). Maintaining participant engagement in digitally delivered group-based treatments is an additional challenge, with therapists perceiving online psychotherapy groups to be less effective, in addition to a lack of specific training for providing online group therapy (Gullo et al., 2022). The highlighted research primarily focused on DMHI generally and did not identify how the shift to digital delivery affected PPTE-focused treatments specifically. Taken together, these challenges may contribute to the low acceptability of DMHI among clinicians (Fluety and Almond, 2020), even with the knowledge that DMHI may provide many unique benefits. Further, until the pandemic, many healthcare regulatory bodies did not provide specific guidelines regarding the utilization of DMHI; as such, many healthcare providers reported a lack of clarity with provision of PPTE-focused therapies in the digital environment, given the lack of regulatory direction.

Most current research into the use of DMHI for the purpose of trauma-focused treatment centers almost exclusively on military and Veteran populations in the United States. This geographic focus highlights a critical lack of research specifically examining the experiences of Canadian PSP, military members, and Veterans who have received psychotherapies utilized for trauma-affected populations. Differences in social and cultural upbringing, differing experiences with the healthcare system, and having comparatively less access to large urban centers compared to those living in the United States may impact how Canadians interact with and experience DMHI. Further research is needed to better understand if and how psychotherapies utilized for trauma-affected populations can be utilized for Canadians seeking treatment following exposure to PPTE.

The aim of this study was to investigate the perceptions of digitally delivered psychotherapies in use for trauma-affected populations, as reported by Canadian military members, Veterans, and PSP who have undergone these interventions. Additionally, the study sought insights from mental health clinicians who provided such interventions to these populations. To understand the strengths and weaknesses of these interventions, the methodological approach included surveys adapted from the Alberta Quality Matrix for Health (AQMH) and the Unified Theory of Acceptance and Use of Technology (UTAUT), as well as semi-structured interviews and focus groups.

## 2. Methods

The current study used an embedded mixed-methods design (Barkin, Schlundt, & Smith, 2013) in a community-engaged research setting (Esmail, Moor, & Rein, 2015). Client and clinician study participants completed a battery of surveys administered using Research Electronic Data Capture (REDCap), a secure web application used for building and managing online surveys and databases (Harris et al., 2009; Harris et al., 2019). Both the client and clinician versions of the survey included demographics, and a customized version of the AQMH and UTAUT survey. Participants were invited to participate in either a 30–60-minute semi-structured interview or focus group, conducted over Zoom, to explore further their perspectives on psychotherapies utilized for trauma-affected populations.

Study data were stored on the REDCap server and a dedicated, encrypted, and password-protected research drive hosted by the Faculty of Rehabilitation Medicine at the University of Alberta. Ethics approval was obtained from the University of Alberta’s Health Research Ethics Board (Pro00109065).

### 2.1 Participant Inclusion and Exclusion Criteria

#### Clients

Currently serving military members, Veterans, and PSP who were receiving or who have received digitally delivered trauma therapy from a mental health clinician in Canada, either through an Operational Stress Injury Clinic (military member and Veteran clients only) or private provider, were recruited. All client participants had a current or prior primary diagnosis of PTSD and/or a trauma-related mental health disorder, which may have stemmed from operational injuries or past adverse experiences (e.g., adverse childhood events). Individuals who were under 18 years of age, and/or unable to provide informed written consent, and/or not fluent in English were excluded from the study.

##### Mental Health Clinicians

Clinician participants included multidisciplinary mental health clinicians and providers (e.g., psychiatrists, psychologists, social workers, and nurse practitioners) practicing in Canada who provided psychotherapies utilized for trauma-affected populations to Canadian military members, Veterans, and PSP. Participants who were not able to provide informed written consent and/or were not fluent in English were also excluded.

### 2.2 Recruitment and Data Collection

Word of mouth, snowball, and purposeful sampling strategies were used to recruit participants from partner and non-partner mental health clinics. Clinics were provided with study recruitment materials and information, which were passed onto potential client and clinician participants. Interested participants who completed a consent to contact form over REDCap were contacted by a member of the research team by telephone or email to discuss the study, determine eligibility, and assess their willingness to participate voluntarily. Recruitment took place between January 2022 to March 2023.

A link to a REDCap webpage was shared with eligible and interested individuals to access and complete the informed consent form. Participants who provided informed consent completed survey measures and/or a semi-structured interview or focus group. Surveys were iteratively developed based on the AQMH (Health Quality Council of Alberta, 2017) and UTAUT (Venkatesh et al., 2003) by the research team. These measures were crafted to maximally align survey language with each AQMH and UTAUT dimension, integrating principles of equity, diversity, and inclusion, minimizing survey burden, and maximizing data quality, collection, and analyses. Data collection took place from February 2022 to May 2023.

### 2.3 Tools and Measures

#### Alberta Quality Matrix for Health Survey (AQMH)

The AQMH was created in 2005 based on the work of *Crossing the Quality Chasm: A New Health System for the 21st Century* (Institute of Medicine (IOM), 2001). The AQMH can be used to organize information about complex health systems for analyses and create awareness of quality in service delivery. The AQMH has the following two components: (1) dimensions of quality, which focuses on aspects of the patient and client experience; and (2) areas of need, which divides services provided by the health system into four distinct but related categories (being healthy, getting better, living with illness or disability, and end of life). The components are each considered across the following six dimensions:

1. Acceptability: health services are respectful and responsive to user needs, preferences, and expectations.
2. Accessibility: health services are obtained in the most suitable setting at a reasonable time and distance (for this paper, accessibility is analogous to access to care and does not refer to designing services for people with disabilities).
3. Appropriateness: health services are relevant to user needs and are based on accepted or evidence-based practice.
4. Effectiveness: health services are based on scientific knowledge to achieve desired outcomes.
5. Efficiency: resources are optimally used in achieving desired outcomes.
6. Safety: services mitigate risks to avoid unintended or harmful results.

The research team iteratively developed a survey based on the AQMH and previous literature (see Appendix 1 for a copy of the AQMH and the survey adapted from the AQMH). The AQMH survey consisted of 10 questions, scored on a 7-point Likert-type scale from 1 (*strongly disagree*) to 7 (*strongly agree*). The conversion allowed participants to rate the quality-of-service of in-person and digital delivery of trauma therapies along AHQM dimensions, using one question to assess each of the following ten criteria: ease of use, convenience, acceptability, practicality, accessibility, appropriateness, effectiveness, efficiency, safety, and fit.

#### Unified Theory of Acceptance and Use of Technology (UTAUT) Survey

The UTAUT model was developed by Venkatesh et al. (2003) as a synthesis of eight technology acceptance models. The UTAUT is designed to assess the acceptance of new technology and may explain up to 70% of the variance in intention to use technologies (Venkatesh et al., 2003). The UTAUT has well established construct and content validity. The six factors influencing technology use as measured by the UTAUT include:

1. Performance Expectancy: assesses whether the participant believed that digital delivery would improve the performance of the job they were trying to complete. If belief in digitally delivered psychotherapies utilized for trauma-affected populations was high, participants would be more likely to use the technology.
2. Effort Expectancy: the degree of ease associated with using digital delivery. If participants perceived digitally delivered psychotherapies utilized for trauma-affected populations to have low Effort Expectancy, it would be expected that they would be more likely to use it.
3. Social Influence: the extent to which individuals surrounding the participant perceived the usefulness of digitally delivered psychotherapies utilized for trauma-affected populations and how much these important other’s influenced participant’s use.
4. Facilitating Conditions: the extent to which conditions, such as organizational and technical infrastructure, surrounding the participant support the use of digitally delivered psychotherapies utilized for trauma-affected populations.
5. Behavioral Intention: the degree to which participants had a conscious plan to utilize digitally delivered psychotherapies utilized for trauma-affected populations. This construct in turn predicts Use Behavior and technology acceptance.
6. Use Behavior: the extent to which participants used digitally delivered psychotherapies utilized for trauma-affected populations.

The research team iteratively developed both client and clinician participant surveys based on the UTAUT content and components, as well as previous literature (see Appendix 1 for a copy of the UTAUT and the surveys adapted from the UTAUT). The UTAUT surveys consisted of 18 questions, scored on a 7-point Likert-type scale from 1 (*strongly disagree*) to 7 (*strongly agree*). Each of the 6 UTAUT constructs was measured individually, with 3 questions asked per construct. All construct scores were then combined to assess the overall useability of the technology used for digitally delivered trauma therapy (Venkatesh et al., 2003).

#### Interviews and Focus Groups

Qualitative data, collected via 30–60-minute semi-structured solo interviews (n=7) or focus groups (n=2) conducted and recorded over Zoom (Archibald et al., 2019), served to further explore participant perspectives on psychotherapies for trauma-affected populations. Separate focus groups were facilitated by members of the study team for both client and clinician participants using a semi-structured interview and focus group guide. Key topics of discussion included the previous and current state of using psychotherapies utilized for trauma-affected populations in the midst of the COVID-19 pandemic; barriers to, facilitators of, and recommendations for the use of psychotherapies utilized for trauma-affected populations to deliver mental health services to military members, Veterans, and PSP; acceptance and methods of delivery for psychotherapies utilized for trauma-affected populations; clinical effectiveness; and needs, including infrastructure and implementation. See Appendix 2 for copies of interview scripts used for client and clinician interviews and focus groups.

Focus groups were purposely heterogeneous with respect to professional representation and experience with using psychotherapies utilized for trauma-affected populations. This design allowed for broad crosstalk and pollination of complementary and alternative ideas, experiences, and conversation, resulting in rich comprehensive data such that sufficient information power was reached (Malterud, Siersma, & Guassora, 2016).

### 2.4 Quantitative Data Analysis

After completion of data collection, all data were de-identified prior to analysis. De-identified survey data were analyzed using IBM SPSS Statistics software (Version 28.0) (IBM Corp., 2021), while NVIVO 13 (2021, R1) was used to facilitate qualitative analysis of de-identified interview and focus group data.

Descriptive statistics were calculated for each survey variable for client and clinician participants. Non-parametric analyses were conducted due to the relatively small sample size of each participant group. Paired-samples Wilcoxon signed-rank tests were used to assess within-subject differences (p≤ 0.05) in median AQMH survey dimension scores between digitally delivered and in-person psychotherapies. One-sample Wilcoxon signed-rank tests were also used to assess statistically significant within-subject differences (p≤ 0.05) between observed and reference median UTAUT survey dimension scores, where the reference score was 12 (i.e., neutral score, based on the sum of three total questions per UTAUT dimension asked on a Likert scale from 1 to 7, where 4 is the median score for each question). In total, 20 tests were conducted for the AQMH survey results (10 for clients and 10 for clinicians) and 12 tests were conducted for the UTAUT survey results (6 for clients and 6 for clinicians). To address multiple comparisons across the 32 statistical tests, the Benjamini-Hochberg procedure was used to control the False Discovery Rate (FDR). A summary of statistical tests can be found in Appendix 3.

### 2.5 Qualitative Data Analysis

Video-recorded interviews and focus groups were transcribed using Adobe Premiere Pro, with transcription accuracy checked by a research team member (SY or RW), and thematically analyzed both deductively and inductively following an iterative process (Braun & Clarke, 2006). Deductively, initial codes were developed based on interview and focus group topics and study objectives. Inductive coding involved identifying themes that emerged from collected data. Coding for each interview and focus group was independently conducted by two research team members (SY, RW) after which a senior researcher (SBP) reviewed and refined the codes. These were then combined and tabulated into preliminary themes. Analysis of preliminary themes by the larger research team followed, with differences being resolved through discussion. A proposed thematic theory then underwent collective analysis, where preliminary themes were modified, and key quotes isolated to illustrate the selected themes. The final thematic narrative was then prepared. The Standard for Reporting Qualitative Research was used to guide the reporting process (O’Brien et al., 2014).

## 3. Results

### 3.1 Demographics

#### Client Participants

Eleven Canadian military members, Veterans, and PSP completed survey measures. Client participants had an average age of 50±10.5 years (range: 34 to 61 years old) and self-identified as female (*n*=3), male (*n*=8), as Caucasian (n=9) and Aboriginal/Metis (n=2). No participants identified as transgender or gender diverse. Our participants self-reported being in the CAF (n=7), police force (n=2), or as a paramedic (n=1), or a correctional worker (n=1), with an average length of service of 21±9.6 years (range: 5 to 38 years). Most participants (n=8) reported having received some form of psychotherapeutic treatment, including CBT (n=5), CPT (n=1), EMDR (n=3), and PE (n=2), for their trauma prior to participating in the current study. Our data, however, did not allow us to distinguish between those who had experience receiving in-person trauma therapy from those who had not received non-trauma specific therapy previously. Four participants had received digitally delivered therapies prior to engaging in the study, while 7 were receiving digitally delivered therapies for the first-time during study participation. All participants had received psychotherapies utilized for trauma-affected populations via video conferencing, with some reporting having received additional services via telephone (n=3).

Four client participants took part in a semi-structured interview (comprising: 2 females, 2 males; 3 Caucasian, 1 Aboriginal/Metis). Participants self-reported being in the CAF (n=2) or police force (n=2) with an average length of service of 19±11.9 years (range: 5 to 31 years). Two participants reported having received some form of psychotherapeutic treatment, including CBT (n=2), EMDR (n=2), and PE (n=1), for their trauma prior to participating in the current study. One participant had received digitally delivered therapies prior to engaging in the study, while three were receiving digitally delivered therapies for the first-time during study participation.

#### Mental Health Clinicians

Twelve Canadian mental health clinicians completed survey measures. Clinician participants self-identified as female (*n*=9), male (*n*=3), Caucasian (n=12), with an average age of 46±8.3 years (range: 33 to 58 years). No participants identified as transgender or gender diverse. Clinician participants held a variety of clinical roles, including registered psychologist (*n*=4), social worker (*n*=4), psychiatrist (*n*=2), and mental health therapist (*n*=2). Workplaces were highly varied and included working at a community mental health clinic (*n*=5), the provincial health authority (*n*=3), private practice (*n*=3), or a regional health service (*n*=1). On average, the clinicians had 14±6.3 years (range: 5 to 25 years) of clinical experience and 9±5.3 years (range: 3 to 16 years) of experience specifically providing trauma therapies. Clinician participants reported using several therapeutic modalities within the digital environment, including Cognitive Behavioral Therapy (*n*=7) (it was unknown if CBT interventions were specifically trauma-focused), Cognitive Processing Therapy (*n*=6), Eye Movement Desensitization and Reprocessing (*n*=10), Prolonged Exposure (*n*=4), Somatic therapy (*n*=2), Mindfulness/Self-Compassion (*n=*3), Dialectical Behavior Therapy based therapy (*n=*5), Acceptance and Commitment Therapy (*n=*2), and Neurotherapy (*n=*1). These modalities were used to treat symptoms related to diverse psychological traumas, including, but not limited to, operational/occupational injuries, adverse childhood events, and sexual abuse/rape. Several participating clinicians (n=5) had not used any form of digital delivery prior to the COVID-19 pandemic. At the time of study participation, six clinicians exclusively provided digitally delivered care, while the remaining six clinicians provided a combination of digitally delivered and in-person care.

In total, 24 clinician participants took part in a semi-structured interview or focus group. Three clinicians participated in a semi-structured interview (2 female, 1 male), while the remaining 21 participated in one of two focus groups (focus group 1: 19 females, 2 males; focus group 2: 12 females, 2 males); there were no differences in participant sex and gender identity. Participating clinician workplaces included a community mental health clinic (*n*=12), within the provincial health authority (*n*=10), or private practice (*n*=2). On average, the clinicians had 13±9.7 years (range: 0 to 40 years) of clinical experience and 6±4.4 years (range: 0 to 15 years) of experience specifically providing trauma therapies.

### 3.2 Survey Results

#### 3.2.1 Client Participants Survey Results

##### Alberta Quality Matrix for Health (AQMH) Survey

Participants rated convenience (*p* = 0.011), practicality (*p* = 0.011), accessibility (*p* = 0.012), and efficiency (*p* = 0.014) scores statistically significantly higher for the digital delivery of trauma therapy compared to in-person delivery. There were no statistically significant differences in client participant ratings for the ease of use, acceptability, appropriateness, effectiveness, safety, and fit dimensions between digital and in-person delivered trauma therapies (Figure 1, Table 1).

**Figure 1.**
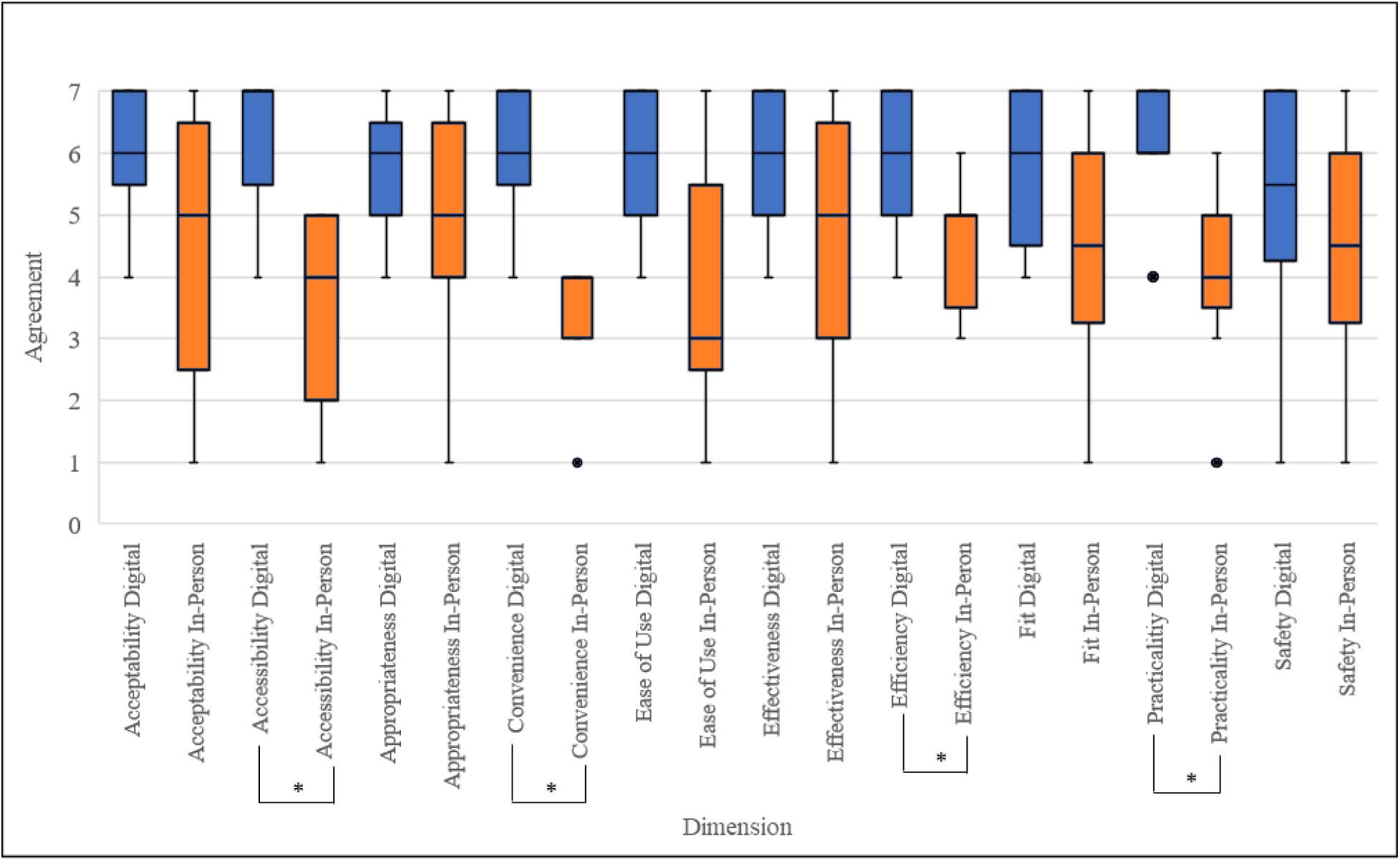
Box and whisker plots indicating client participant (n=11) median AQMH survey scores, first and third quartiles, and minimum and maximum scores. *=Significant difference (p<0.05) between median scores for digital delivery vs in-person therapy based on paired sample Wilcoxon signed-rank test, corrected for multiple comparisons. Blue refers to digital delivery; orange refers to in-person delivery. ● indicates outlier.

**Table 1.**
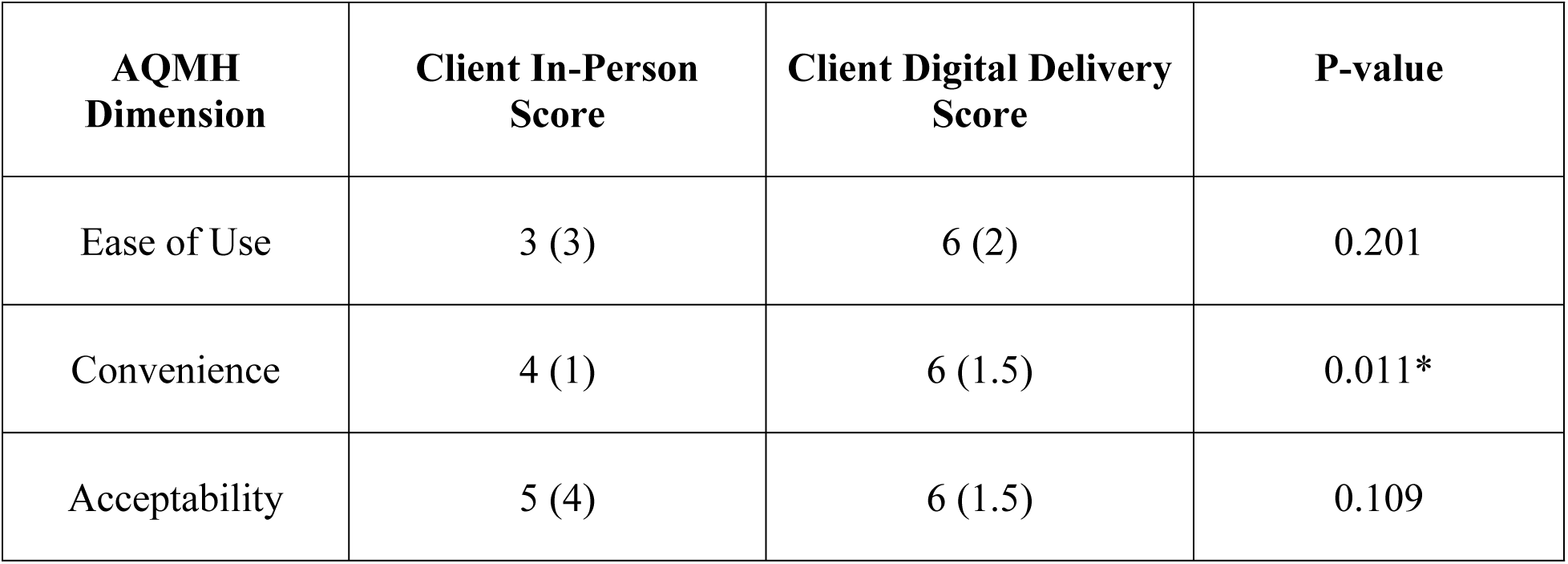
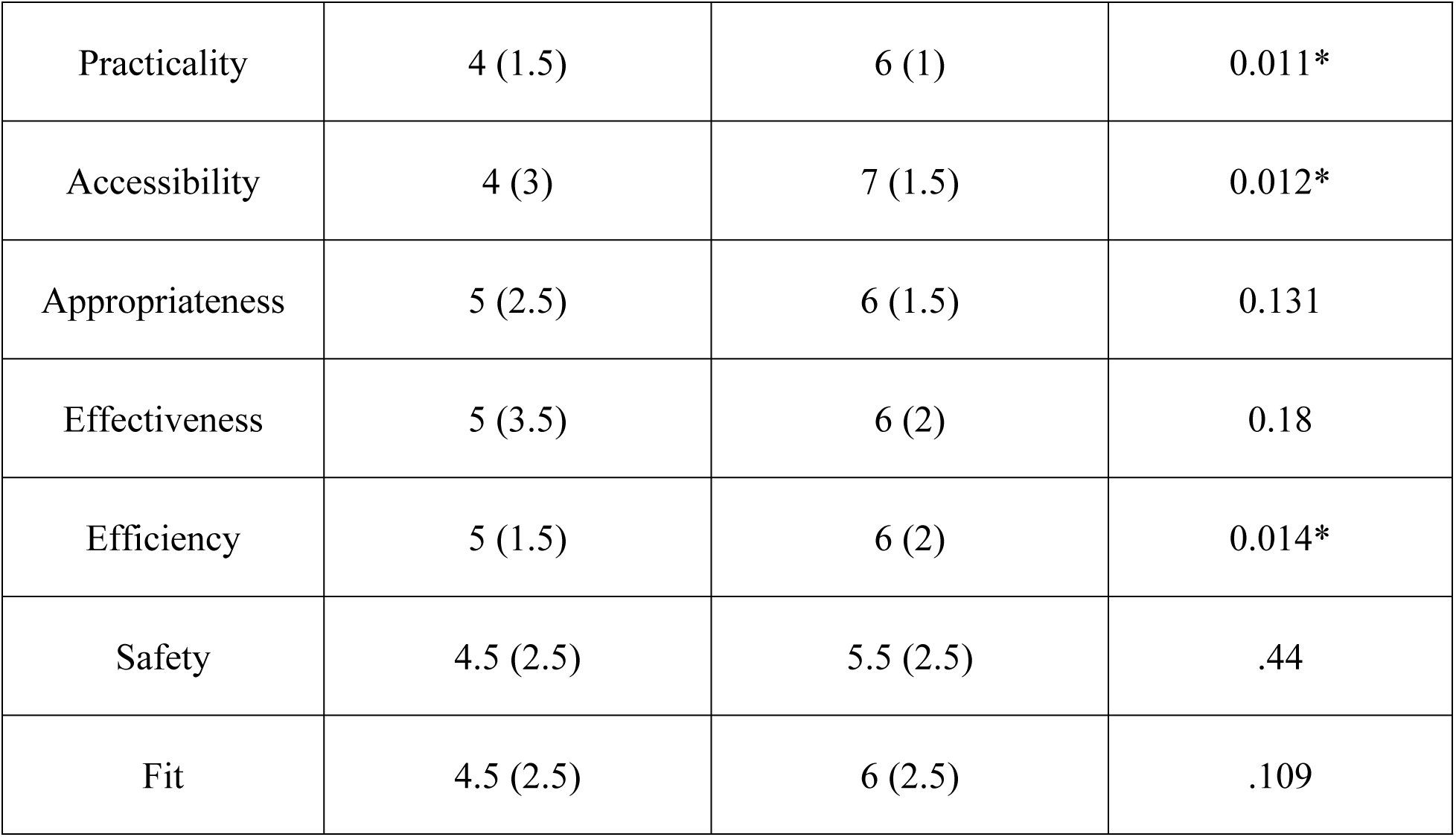
Client participant (n=11) median AQMH survey scores with interquartile ranges. *=Significant difference (p<0.05) between median scores for digital delivery vs in-person therapy based on paired sample Wilcoxon signed-rank test, corrected for multiple comparisons.

##### Unified Theory of Acceptance and Use of Technology (UTAUT) Survey

All client participants agreed that digitally delivered trauma therapy services were a viable alternative to in-person delivery. Client participants indicated they somewhat agreed, agreed, or strongly agreed with the Effort Expectancy (score: 16/21), Performance Expectancy (score: 17/21), Behavioral Intention (score: 18/21), and Use Behavior (score: 15/21) constructs. Analysis revealed that only Performance Expectancy (p=0.011) scores were significantly different compared to expected median scores. Client participants also indicated that they neither agreed nor disagreed with the Social Influence (score: 12/21) and Facilitating Conditions (score: 13/21) constructs (Figure 2, Table 2).

**Figure 2.**
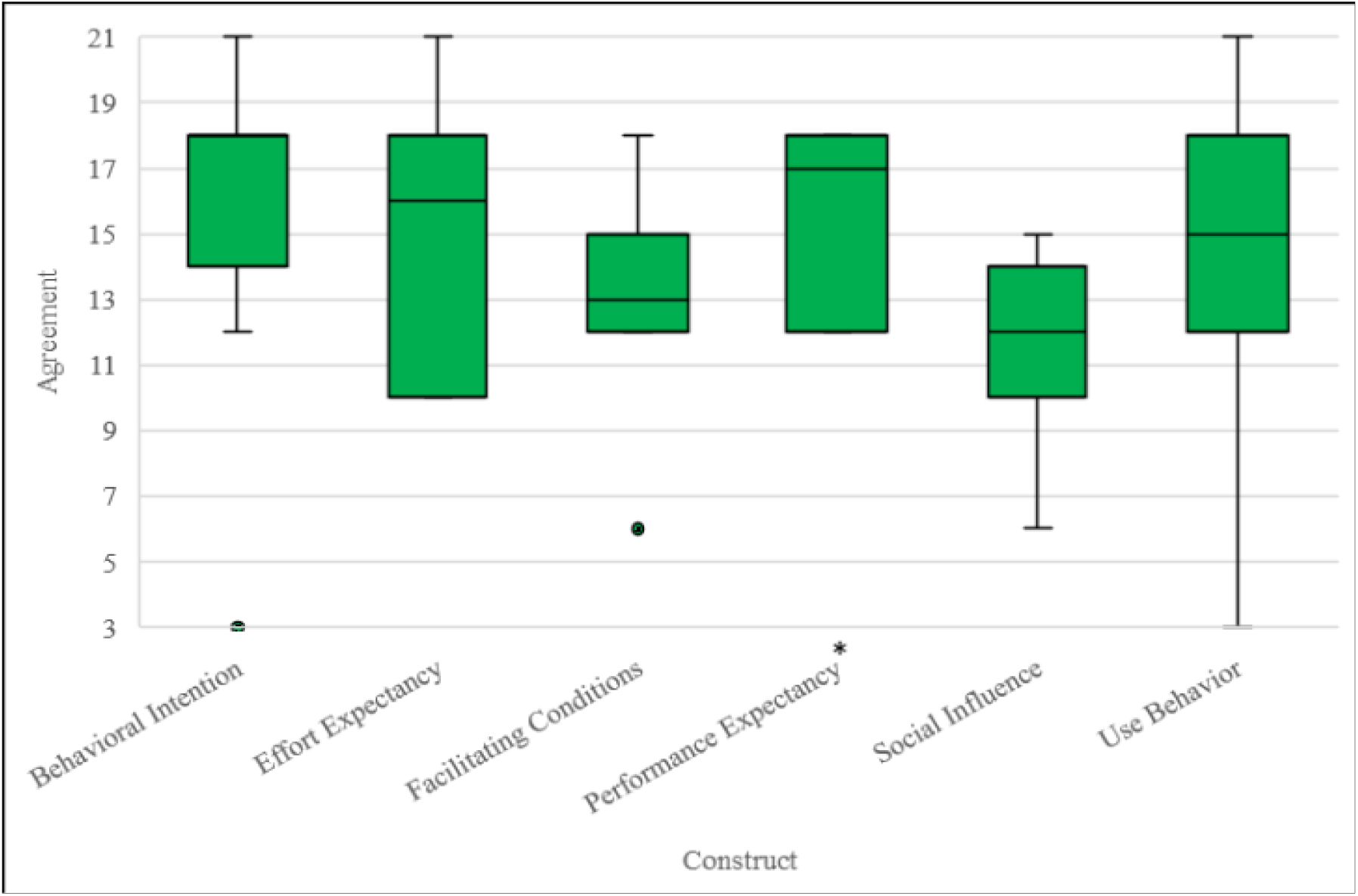
Box and whisker plots indicating client participant (n=11) median UTAUT construct scores, first and third quartile, and minimum and maximum scores. *=Significant difference (p<0.05) between median score and a reference score of 12 (total score of three questions asked based on Likert scale 1-7) based on one-sample Wilcoxon signed-rank test, corrected for multiple comparisons. ● indicates outlier.

**Table 2.** Client participant (n=11) median UTAUT survey scores with interquartile ranges. *=Significant difference (p<0.05) between median score and a reference score of 12 (total score of three questions asked based on Likert scale 1-7) based on one-sample Wilcoxon signed-rank test, corrected for multiple comparisons.

| UTAUT Dimension | Client Median Agreement Score (interquartile range) | P-value |
| --- | --- | --- |
| Performance Expectancy | 17 (6) | 0.011* |
| Effort Expectancy | 16 (8) | 0.032 |
| Social Influence | 12 (4) | 0.67 |
| Facilitating Conditions | 13 (3) | 0.368 |
| Behavioral Intention | 18 (4) | 0.057 |
| Use Behavior | 15 (6) | 0.182 |

#### 3.2.2 Mental Health Clinician Participant Survey Results

##### Alberta Quality Matrix for Health (AQMH) Survey

There were no statistically significant differences between Clinician participant AQMH scores for digitally delivered vs. in-person therapy for all 10 AQMH dimensions: ease of use, convenience, acceptability, accessibility, practicality, appropriateness, effectiveness, efficiency, safety, and fit (Figure 3, Table 3).

**Figure 3.**
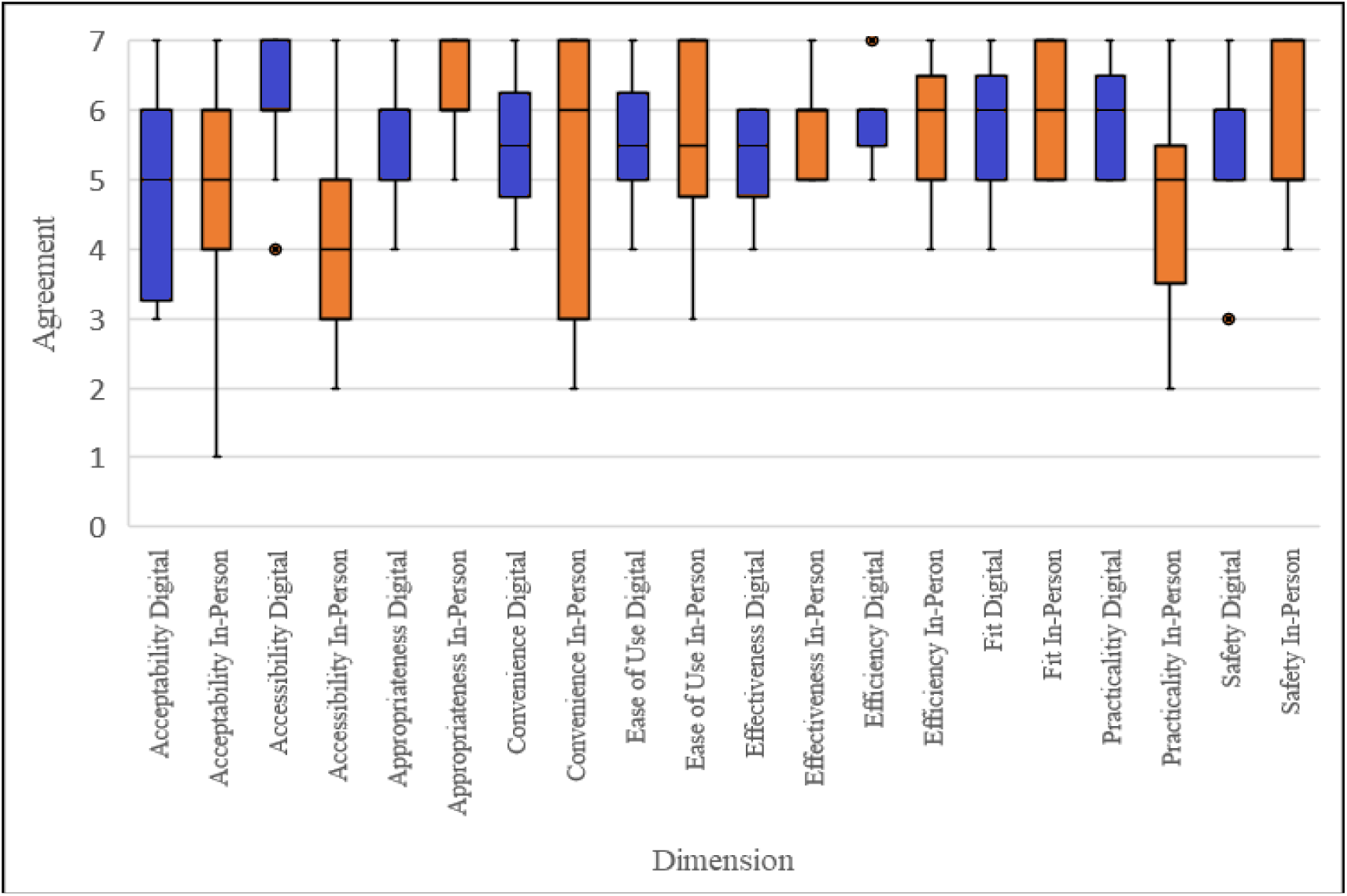
Box and whisker plots indicating clinician participant (n=12) median AQMH survey scores, first and third quartiles, and minimum and maximum scores. *=Significant difference (p<0.05) between median scores for digital delivery vs in-person therapy based on paired sample Wilcoxon signed-rank test, corrected for multiple comparisons. Blue refers to digital delivery; orange refers to in-person delivery. ● indicates outlier.

**Table 3.** Clinician participant (n=12) median AQMH survey scores with interquartile ranges. *=Significant difference (p<0.05) between median scores for digital delivery vs in-person therapy based on paired sample Wilcoxon signed-rank test, corrected for multiple comparisons.

| AQMH Dimension | Clinician In-Person Score | Clinician Digital Delivery Score | P-value |
| --- | --- | --- | --- |
| Ease of Use | 5.5 (2.5) | 5.5 (1) | 0.943 |
| Convenience | 5 (3) | 5.5 (2.5) | 0.681 |
| Acceptability | 5 (1.75) | 5 (2.5) | 0.952 |
| Practicality | 5 (2.5) | 6 (1.75) | 0.114 |
| Accessibility | 4 (2.75) | 6 (1) | 0.020 |
| Appropriateness | 6 (1.75) | 6 (1) | 0.046 |
| Effectiveness | 6 (1) | 6 (1) | 0.194 |
| Efficiency | 6 (1) | 6 (0.75) | 0.785 |
| Safety | 5 (2) | 6 (1) | 0.492 |
| Fit | 6 (2) | 6 (1.75) | 0.395 |

##### Unified Theory of Acceptance and Use of Technology (UTAUT) Survey

Overall, clinician participants strongly agreed that digitally delivered psychotherapies utilized for trauma-affected populations were a viable option when compared to in-person trauma therapies. Clinician participants indicated slight agreement, agreement, or strong agreement with the Effort Expectancy (18/21), Performance Expectancy (18.5/21), Behavioral Intention (21/21), Use Behavior (18.5/21), and Social Influence (16/21) constructs. Analysis revealed that Behavioral Intention (p=0.002), Performance Expectancy (p=0.003), Use Behavior (p=0.004), and Effort Expectancy (p=0.012) scores were significantly different compared to expected median score of 4 (on Likert scale 1-7), surviving FDR multiple comparison correction. Clinician participants indicated that they neither agreed nor disagreed with the Facilitating Conditions (15/21) construct (Figure 4, Table 4).

**Figure 4.**
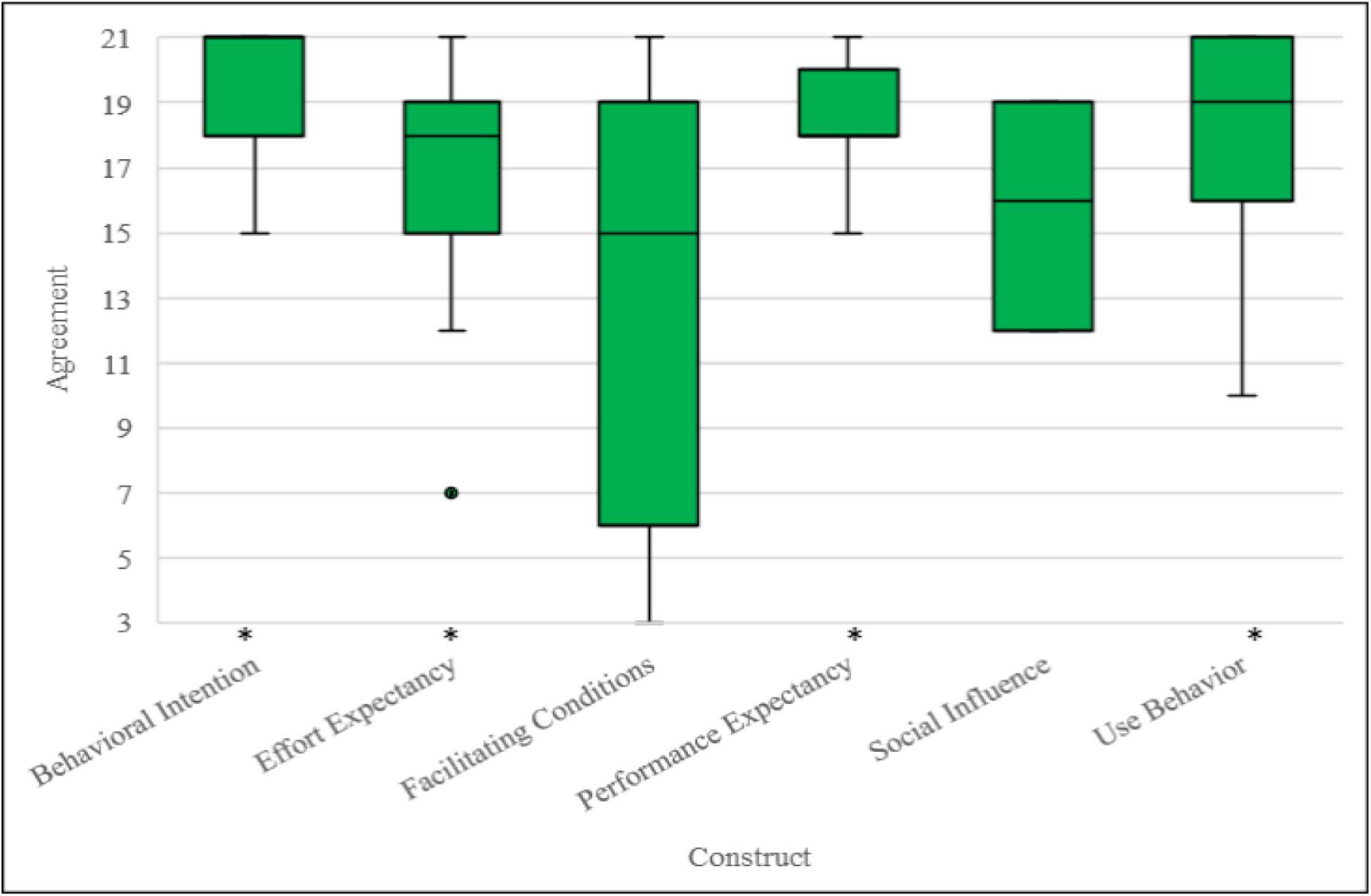
Box and whisker plots indicating clinician participant (n=12) median UTAUT construct scores, first and third quartiles, and minimum and maximum scores. *=Significant difference (p<0.05) between median score and a reference score of 12 (total score of three questions asked based on Likert scale 1-7) based on one-sample Wilcoxon signed-rank test, corrected for multiple comparisons. ● indicates outlier.

**Table 4.** Clinician participant (n=12) median UTAUT survey scores with interquartile ranges. *=Significant difference (p<0.05) between median score and a reference score of 12 (total score of three questions asked based on Likert scale 1-7) based on one-sample Wilcoxon signed-rank test, corrected for multiple comparisons.

| UTAUT Dimension | Clinician Median Agreement Score (interquartile range) | P-value |
| --- | --- | --- |
| Performance Expectancy | 18.5 (2) | 0.003* |
| Effort Expectancy | 18 (4) | 0.012* |
| Social Influence | 16 (7) | .017 |
| Facilitating Conditions | 15 (13) | 0.593 |
| Behavioral Intention | 21 (3) | 0.002* |
| Use Behavior | 18.5 (5) | 0.004* |

### 3.3 Interview Results

Thematic analysis of the interview data isolated five main themes with regard to psychotherapies utilized for trauma-affected populations: (1) Digital delivery similar to in-person care; (2) Unique benefits of digital delivery; (3) Digital delivery and reduced emotion; (4) Concerns regarding digital delivery; and (5) Future directions and recommendations.

#### Theme 1: Digital delivery similar to in-person care (Table 5)

Client participants (4/4) felt very comfortable receiving digitally delivered psychotherapies utilized for trauma-affected populations. This comfort reportedly stemmed from their familiarity using digital platforms such as Zoom in their occupations and daily lives, as these platforms saw increased usage following the onset of the COVID-19 pandemic (Karl, Peluchette, and Aghakhani, 2022).

**Table 5.** Digital Delivery Similar to In-person Care.

| Sub Theme | Description and Supportive Quotes |
| --- | --- |
| 5.1 Therapy Effectiveness | Certain psychotherapeutic modalities translated very well to digital delivery. This left clinician participants to perceive that digital delivery conferred similar quality of care as in-person delivery.<br>*“I’ve had people who’ve done full courses of treatment doing EMDR online, I’ve actually never met them in person. It was beautiful, very effective.” [Clinician participant 3]* |
| 5.2 Connection with Therapist | Being comfortable and familiar with working in digital environments made client participants feel more ready to engage with digitally delivered psychotherapies utilized for trauma-affected populations, which in turn aided in developing a strong therapeutic alliance with their clinician.<br>*“I didn’t because I had used it. I’ve been using it since the beginning of COVID with my work that I was doing. [...] I felt just as close to the virtual therapist as the in-person one. I did feel a real sense of bonding with them and feeling as if I could disclose things and talk to them[.] I didn’t find it different in that sense[.] I made a very strong connection with the therapist very quickly. She was excellent. Very experienced, and I felt very comfortable with her.” [Client participant 4]* |

Clinician participants (18/24) also shared that they felt that psychotherapies utilized for trauma-affected populations provided similar quality of care as in-person delivery. In particular, clinician participants shared that patient assessments and certain treatment modalities, including EMDR and CBT, were easy to adapt to the digital environment and resulted in successful administration of digitally delivered treatment with no discernable differences to in-person delivery.

#### Theme 2: Unique benefits of digital delivery (Table 6)

Client and clinician participants all agreed that the most crucial benefit of digital delivery was that it increased the accessibility of psychotherapies utilized for trauma-affected populations. Having digital therapy sessions allowed for more flexible scheduling, allowing client participants to attend sessions without sacrificing other responsibilities (e.g., not missing work or childcare responsibilities) and clinician participants to reach clients living in remote locations.

**Table 6.**
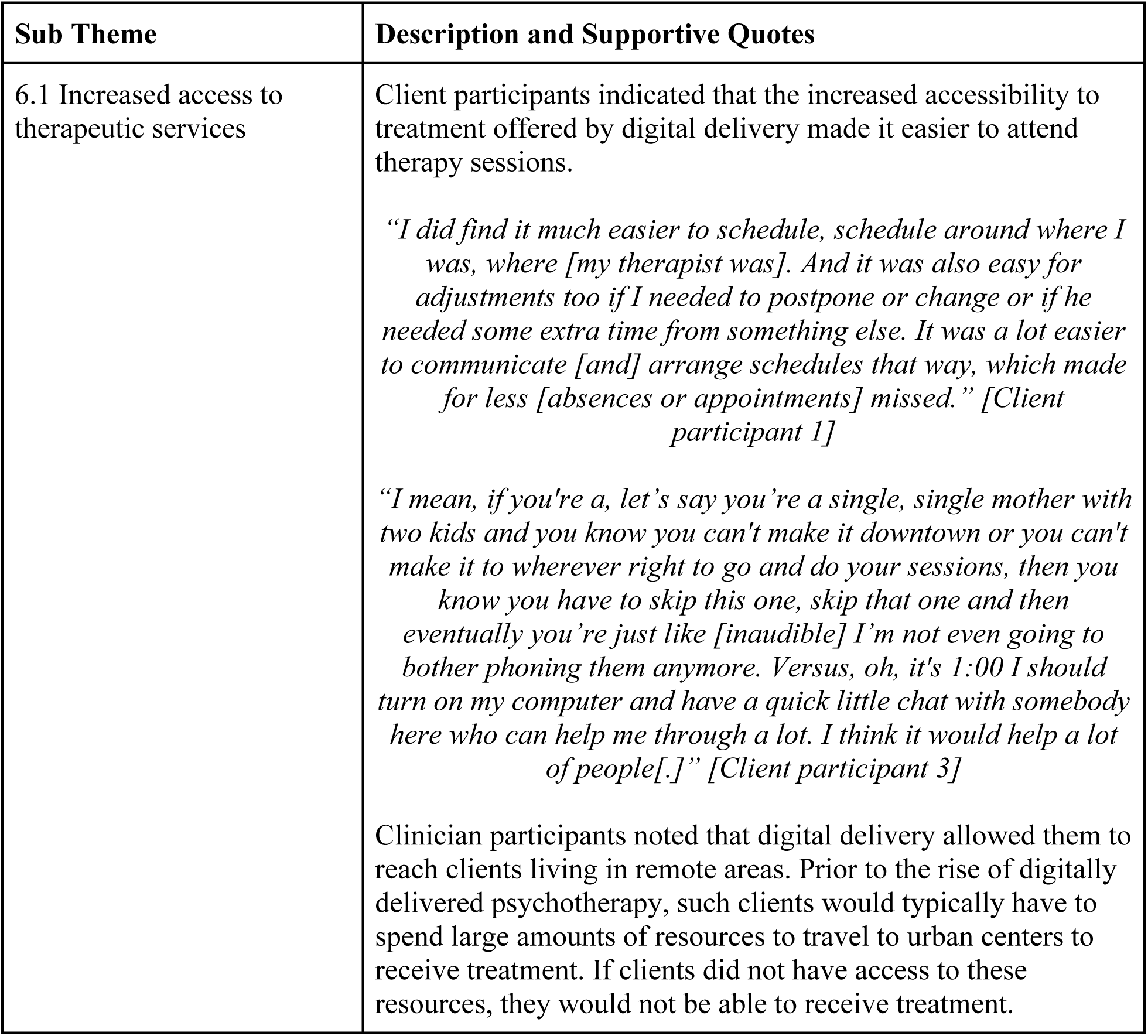
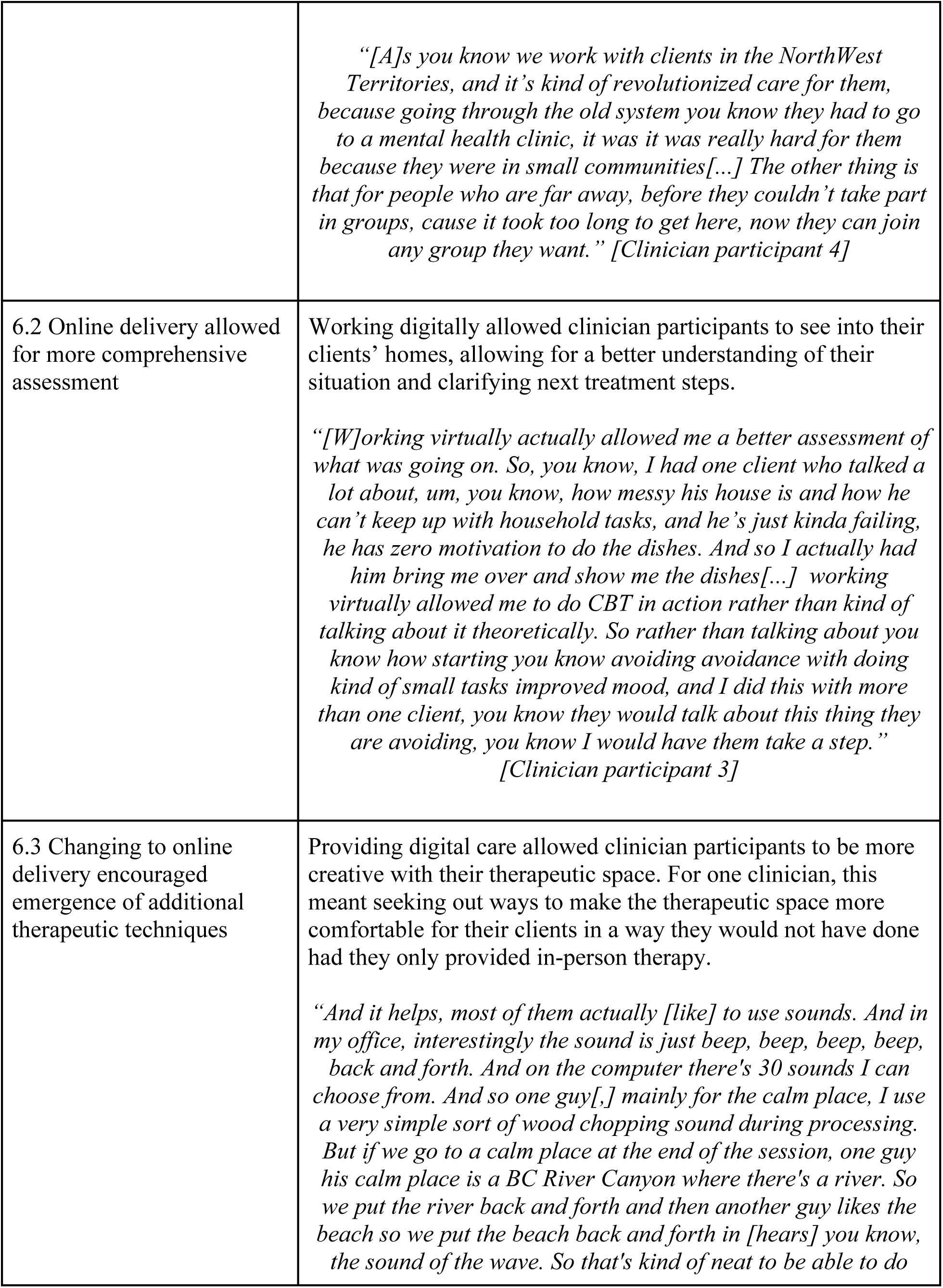
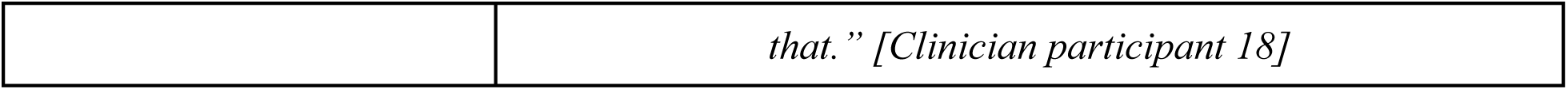
Unique Benefits of Digital Delivery.

Some clinician participants (8/24) revealed that digital delivery aided in their therapeutic duties in creative ways not thought of when delivered in-person care. This led to clinicians conducting more comprehensive assessments of clients or creating more comfortable therapeutic environments for clients.

#### Theme 3: Digital delivery and reduced emotion (Table 7)

For 2 client participants, working in a digital environment left them experiencing fewer emotions compared to when they had received in-person care. One participant experienced less stress receiving digitally delivered care, while the other experienced fewer emotions in general working in the digital environment.

**Table 7.**
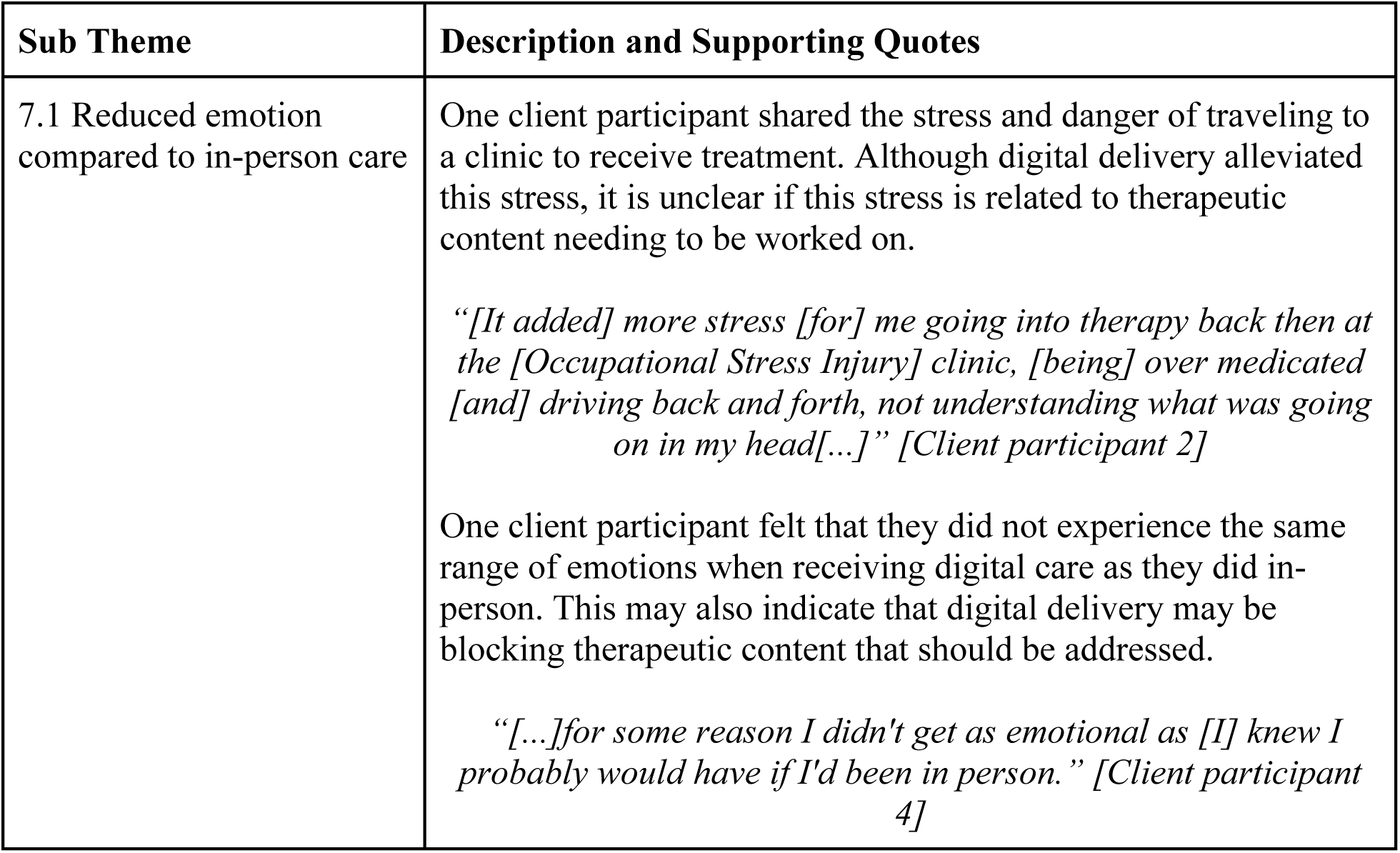
Digital Delivery and Reduced Emotion.

| Sub Theme | Description and Supporting Quotes |
| --- | --- |
| 7.1 Reduced emotion compared to in-person care | One client participant shared the stress and danger of traveling to a clinic to receive treatment. Although digital delivery alleviated this stress, it is unclear if this stress is related to therapeutic content needing to be worked on.<br>*“[It added] more stress [for] me going into therapy back then at the [Occupational Stress Injury] clinic, [being] over medicated [and] driving back and forth, not understanding what was going on in my head[...].” [Client participant 2]*<br>One client participant felt that they did not experience the same range of emotions when receiving digital care as they did in-person. This may also indicate that digital delivery may be blocking therapeutic content that should be addressed.<br>*“[...]for some reason I didn't get as emotional as [I] knew I probably would have if I'd been in person.” [Client participant 4]* |

#### Theme 4: Concerns regarding digital delivery (Table 8)

Some client participants (2/4) found that attending digital sessions could be somewhat resource intensive, with attendance typically requiring a general knowledge of using digital platforms, having access to a working computer, stable high speed internet connection, web camera with clear picture, a quiet and private space to attend the session, and access to a support person nearby and available in the event of an adverse emotional response. Client participants (3/4) also raised concerns over family members or significant others overhearing their trauma therapy sessions, raising questions regarding the security and privacy of digitally delivered services. Such challenges left some client participants (2/4) feeling disconnected from their clinician and the therapeutic experience in general, leading to less effective treatment and a less intimate therapeutic relationship. For these participants, this left the impression that psychotherapies utilized for trauma-affected populations may not be an adequate replacement for in-person delivery for some patients or contexts.

**Table 8.** Concerns Regarding Digital Delivery.

| Sub Theme | Description and Supporting Quotes |
| --- | --- |
| 8.1 Privacy | Client participants felt uncomfortable at the idea that other people could hear their digital therapy sessions.<br>*“Yes, so that was a little awkward because, you know, if somebody was in my house and I had to go to a different room, there was always the thought of “Can they hear me? Are they listening?” I mean, generally I was not in a house of somebody I couldn't trust anyways. But, you know, those are private sessions that you want to keep to yourself.” [Client participant 1]*<br>*“There [are] certain things about [my] experiences that I would prefer my family not to know about[,] that I keep to myself, right? You know, [my] five year old is sitting on the couch right now while we're doing this. So, you know, that's a little bit of a negative.” [Client participant 3]*<br>Clinician participants felt a lack of control over the therapeutic space when working digitally, affecting their ability to provide care.<br>*“I could add that I have significantly less control, if not no control, over the confidential therapeutic space, at least on the end of the client in virtual therapy[...] [Clinician participant 2]* |
| 8.2 Safety | Clinician participants shared concerns about client safety. In particular, clinician participants were unsure of how to respond to dangerous relationships their clients were a part of or adverse events their clients would experience when working digitally.<br>*“If a client is in a relationship that has violence or has a history of domestic violence[,] that has been something that's come up for me[,] and I've had to kind of check in with the client every time, is your partner home, are they able to hear it hear in and listen in on this, and I was able to sort of contract with the client around having sessions when the partner was not home.” [Clinician participant 2]*<br>*“[There] sort of [has to be] concerns for safety. How do I help my client that's having an over-the-top reaction or* |
|  | *different reactions if there's something[?] [Y]ou're not there to deal with it directly, you know, especially then if they cut the mic or whatever, it can [happen] abruptly then you know, and you're like what the hell is happening there? I can't be there. No. So yeah, it doesn't come up very often, hardly at all. But, you know, there's something to be concerned about.”*<br>*[Clinician participant 18]* |
| 8.3 Technological Literacy | Client and clinician participants acknowledged that technological literacy varies widely between individuals, and expressed concern with the lack of support being provided to those not comfortable with using digital platforms. Further, participants noted that it is unclear if digital delivery truly allows for equitable access to care.<br>*“He had to be able to talk to me through[,] to talk me through [going] on the Jane chat thing that we, right? He had to talk me through it over the phone, right, because I'm not[,] I'm not into all this stuff so[...] [I was frustrated because] I hate computers, data, and I was just like, I don't know how to operate it.” [Client participant 3]*<br>*“[A gap that should be addressed is providing] practical training in using Zoom and troubleshooting issues for clinicians who aren't as tech savvy.” [Clinician participant 8]*<br>*“Virtual care operates on the assumption clients have access to technology to support this, not all clients may have the funds and or literacy to access this technology” [Clinician participant 13]* |
| 8.4 Digital Environment Contributing to Emergence of Avoidance Behaviors | Clinician participants expressed concern that the shift to digital delivery was followed by the emergence of avoidant behaviors in their clients. As avoidance (implicit and explicit) may contribute to treatment resistance/poor treatment response, these concerns must be addressed moving forward.<br>*“I've had a few people who are a combination of really avoidant, so [strong] avoidance component of PTSD, and then also tend to have maybe some [obsessive compulsive] personality traits, where they're a combination of, if we're doing CPT for example, um there's kind of a lot of really subtle avoidance, and and often sort of getting into uh rumination in the session rather than focusing on their [stuck point][...]” [Clinician participant 7]* |
|  | *“[W]hat I’m finding is that [clients are] starting to isolate more at home, and not engaging in their community or getting out, and everything has defaulted back to the caregivers in the home, it’s that trust. So I-I really noticed that in a couple of clients who were already highly anxious and highly avoidant. So I don’t think it [the] virtual serves a good purpose in that sense.” [Clinician participant 9]* |

Clinician participants (14/24) indicated that a major barrier to providing digitally delivered psychotherapies for trauma-affected populations was the lack of preparedness stemming from the sudden shift to using digital platforms following the onset of the COVID-19 pandemic. Clinicians shared that they received little to no support to prepare for delivering digital psychotherapies for trauma-affected populations and were tasked with troubleshooting technical issues with very limited support, leading to frustration and discomfort. These challenges greatly impacted many clinicians’ therapeutic ability, disrupting the flow of treatment and leaving them feeling like they were losing control over the therapeutic environment.

Clinician participants also raised concerns that digitally delivered psychotherapies utilized for trauma-affected populations may not provide effective treatment for certain clients. Specifically, clinician participants feared that clients with highly avoidant behavior, dissociative tendencies, complex trauma, or emotional dysregulation may not attain the same benefits from digitally delivered trauma therapy as in-person.

#### Theme 5: Future directions and recommendations (Table 9)

Client and clinician participants indicated several recommendations for integrating digital delivery into psychotherapy care for trauma-affected populations, including increasing the opportunities for and the spread of hybrid care, a combination of digital and in-person therapeutic services.

**Table 9.** Future Directions and Recommendations.

| Sub Theme | Description and Supporting Quotes |
| --- | --- |
| 9.1 Hybrid care | Client and clinician participants shared a common belief that hybrid care would offer a balance between providing the care clients are seeking with the delivery method they prefer.<br>*“Yeah, I think maybe something like you're offering them both and if people are selecting virtual because they can get in quicker, letting them know that it could be transitioned [by] agreement or in particular what the therapist thinks would be best if you could transition into in-person somewhere down the road during the therapy. If, if that's if it looks like it's needed. You wouldn't get absolutely stuck with one if [you] chose it. Similarly, you could switch from in-person to virtual, maybe as you're coming to the end of your therapy.” [Client participant 4]*<br>*“Perhaps like increasing [digital delivery or hybrid care] [...] to provide clients with the care they are seeking.” [Clinician participant 17]* |
| 9.2 Increasing spread | Clinician participants encouraged the wider spread of digital delivery such that more clients would have the opportunity to receive care. |
|  | *“Perhaps like increasing [digital delivery or hybrid care] outside of the Edmonton zone. [...] So if people are interested in these groups, so having it more accessible, we got better in terms of the handouts, having it all prepackaged, everything ready to go before the group because initially we were going between all the sessions, session one, session two and so forth. So I think we're more organized, but improving it [and] having more flexibility.” [Clinician participant 17]*<br>A clinician participant also encouraged other clinicians to be open to providing digital care.<br>*“And I think the other thing is just to, you know, embrace [digital delivery] [and] recognize that there are some really significant benefits to it. You know, try it out and see because I know some people are just really reluctant to even try it, you know, some people never [used] online delivery through [the pandemic]. [Clinician participant 19]* |

Taken together, survey and interview data indicate that clients and clinicians believe that digitally delivered psychotherapies utilized for trauma-affected populations have several unique benefits, including better accessibility to treatment and greater client autonomy, all while offering similar therapeutic outcomes as in-person delivery. Several factors, including technical difficulties inherent in use of digitally delivered treatments and potential security and psychological safety, were concerning for study participants. Further investigation is warranted to address the concerns and the recommendations made by client and clinician participants.

## 4.0 Discussion

The current study provides preliminary evidence of client and clinician support regarding the use of DMHI in the context of providing psychotherapy to military members, Veterans, and PSP who have experienced trauma. Client and clinician study participants reported that digitally delivered psychotherapies utilized for trauma-affected populations appeared to offer similar treatment quality of care as in-person delivery while also improving treatment access.

Our study also provides insight into the implementation of new DMHIs in the Canadian context. Canadian clients and mental health clinicians face many unique challenges, including potentially having to travel long distances from remote regions to urban centers to access evidence-based specialized trauma treatment. Clinicians must provide care that is acceptable and appropriate for multiple diverse populations, including Indigenous groups. This unique perspective was a priority for the research team and highlights the importance of co-designing services, keeping in mind the specific needs, such as treatment accessibility and appropriateness, of clients and clinicians. Further, this study provides an update on previous research in the field (Smith-MacDonald et al., 2020), enabling a better understanding of shifts in attitude and usage of psychotherapies utilized for trauma-affected populations over the COVID-19 pandemic. These findings may play an important role when considering the expansion of DMHI services within the general healthcare system and for delivering care to individuals living in rural or remote communities.

AQMH survey data indicated that client and clinician participants expressed no statistically significant differences in their assessments of service quality between digitally delivered and in-person trauma therapies. This is consistent with previous research suggesting equivalent quality care is attainable via digital or in-person service delivery modalities (Jones et al., 2020, Perri et al., 2021). Client and clinician interview data endorsed the findings of the AQMH survey. In addition, client participants rated the convenience, practicality, accessibility, and efficiency of psychotherapies utilized for trauma-affected populations higher than in-person delivery, corroborating previous research on the unique advantages of DMHI (e.g., increased cost-effectiveness, time savings, and access to therapy compared to in-person care) (Fluety and Almond, 2020; Jones et al., 2020). These dimensions of healthcare appear to be highly valued by clients receiving such therapies. Further research is yet needed to verify our results and better understand if these dimensions of healthcare ought to be prioritized within healthcare programs.

Digital delivery of psychotherapies utilized for trauma-affected populations appeared to be acceptable based on the UTAUT-related questions for client and clinician participants. UTAUT-related questions also indicated that Social Influence and Facilitating Conditions did not appear to be as important to client and clinician participants. This would appear to indicate that client and clinician participants were willing to use digitally delivered psychotherapies despite a lack of perceived support from important others (e.g., family) or the organizations they work for (e.g., mental health clinics). Perhaps this points to client safety and treatment needs overriding client and clinician needs for social and organizational support with regards to using DMHI, including psychotherapies utilized for trauma-affected populations. However, clinicians reported a range of results regarding organizational support, indicating that this factor may be context and clinician dependent. Organizational support therefore requires further study, as it is unclear whether sufficient support is being provided to clinicians to maintain the long-term use of digitally delivered psychotherapies. Finally, although there are moderators known to influence Behavioral Intention and overall technology acceptance, due to the limited sample size, the influence of these moderators was not evaluated.

Client and clinician participants raised some concerns regarding the support provided when receiving or providing digitally delivered psychotherapies utilized for trauma-affected populations during interview and focus group sessions. Specifically, client participants shared concerns regarding accessing the appropriate technology required to attend digitally delivered sessions, while clinician participants expressed frustration stemming from a lack of training and easily accessible technological support. Similar findings were found in a qualitative study conducted in the United States which recommended that, among other factors, providing equitable device distribution and digital literacy training for clients, and adequate technology and support for mental health providers and local behavioral health departments, would be necessary for the successful implementation of DMHI (Zhao et al., 2023). Further research is needed to better understand if these factors should be prioritized when implementing DMHI, including psychotherapies utilized for trauma-affected populations.

Other considerations regarding implementation moving forward are the formation of the therapeutic relationship and management of client distress in the digital environment. Some client participants expressed difficulties in forming a strong therapeutic relationship and emotional connection with their clinician, in line with previous research indicating that individuals who have experienced interpersonal trauma may have difficulties developing a strong therapeutic alliance (Lawson, Skidmore, and Akay-Sullivan, 2020). Such difficulties may, in part, relate to why clinician participants felt less control over the therapeutic environment and worried more about clients experiencing distress when providing digital care. This points to a need to create clinical practice supports and guidelines regarding managing client crises when working in a digital environment. Further, for the safety of mental health clinicians, future guidelines should clarify the legal protections clinicians have when providing DMHI. Finally, it would be useful to further evaluate if there are differences in implementation depending on the context in which a clinician provides clinical services (e.g., is DMHI implementation more likely in a clinic-based setting versus an independent practice).

### Limitations and Future Directions

There were several study limitations which must be acknowledged. First, all recruitment and surveys in the current study were in English, which limited responses from non-English speaking communities. Second, the age and experience of our clinician population may have biased our results. The clinician participants for the current study had an average of 14 years of clinical experience, and the clinician experience providing DMHI services may have been different had our population been younger and less experienced. Similar limitations have been described in previous research (Gullo et al., 2022). Finally, the sample size and the diversity of our study population were relatively limited, which precluded analyzing certain mediating effects. For example, although there are moderators known to influence Behavioral Intention and overall technology acceptance, due to the limited sample size, the influence of these moderators was not evaluated. Similarly, potential gender and sex differences were not explored given the limited sample size.

Future studies with larger sample sizes would be useful to replicate the findings reported here and to explore potential moderating factors, such as differences related to gender, sex, and sexual orientation with regards to the acceptance of digitally delivered care.

### Conclusion

Client and mental health clinician participants shared common perspectives that demonstrated unique benefits and barriers of digitally receiving/providing psychotherapies utilized for trauma-affected populations. Given the high rate of PTSIs within this client base, it is critical that they have access to, and clinicians are able to provide, the highest quality interventions in a secure, cost-effective, and accessible manner.

Our results suggest that digital delivery offers an accessible and practical way for Canadian military members, Veterans, and PSP to receive trauma therapy. Further, client and clinician participants indicated that hybrid care, a mixture of digital and in-person delivery, should be expanded upon in the future to ensure client populations are receiving care through their preferred delivery method. This study adds to the growing body of evidence supporting the use of DMHI in trauma-affected populations. Ultimately, the livelihoods of trauma-affected populations may be directly impacted and improved with the use of DMHI.

## Data Availability

All relevant data are within the manuscript and its Supporting Information files.

## Acknowledgements

Funding support for this project was provided by the following agencies:

- Government of Alberta through the Supporting Psychological Health in First Responders (SPHIFR) grant program;
- Canadian Institutes of Health Research (CIHR) through the Knowledge Synthesis: COVID-19 in Mental Health & Substance Use grant competition.

We at HiMARC would like to thank the Edmonton Operational Stress Injury Clinic for their support and contributions to this project.

